# Diagnostic Indexes of a Rapid IgG/IgM Combined Antibody Test for SARS-CoV-2

**DOI:** 10.1101/2020.03.26.20044883

**Authors:** Liu Ying, Liu Yue-ping, Diao Bo, Ren Feifei, Wang Yue, Ding Jinya, Huang Qianchuan

**Author notes:** The corresponding authors: Ding Jinya or Huang Qianchuan. The authors contributed equally to this work (Liu Ying, Liu Yue-ping).

## Abstract

**Objective:** Coronavirus disease 2019 (COVID-19) has become pandemic in the world. The need for IgG-IgM combined antibody test is booming, but data on diagnostic indexes evaluation was inadequate. The aim of this study was to evaluate diagnostic indexes of a rapid IgG-IgM combined antibody test for SARS-CoV-2.

**Methods:** A total of 179 patients were enrolled. Serum were collected for IgG-IgM combined antibody test and corresponding nasal and pharyngeal swab specimens were collected for SARS-CoV-2 RT-PCR. According to SARS-CoV-2 RT-PCR results, patients under study were categorized as PCR positive group in 90 patients and PCR negative group in 89 patients.

**Results:** 1. Of the 90 PCR positive samples, 77 were tested positive by SARS-CoV-2 IgG-IgM test kit, yielding a sensitivity of 85.6%. Meanwhile, of the 89 PCR negative sample, 8 samples were detected positive, resulting in a specificity of 91%. Positive predictive value, negative predictive value and accuracy of this test kit was 95.1%, 82.7%, and 88.3%, respectively. Kappa efficiency between IgG/IgM test kit and RT-PCR were 0.75. 2. Accuracy in mild/common and severe/critical subgroup were 73.9% and 97.7%, respectively. Accuracy in clinical confirmed, suspected cases and other disease subgroups were 70%, 60%, and 100%, respectively. 3. Patients were further divided into ‘0 - 7’, ‘8 - 15’ and ‘>= 16’ groups according to the time from illness onset to sample collection. Sensitivity, specificity and accuracy in these three groups were 18.8%, 77.8% and 40%; 100%, 50% and 87.5%; 100%, 64.3%, and 93.9, respectively.

**Conclusion:** The sensitivity and specificity of this ease-of-use IgG/IgM combined test kit were adequate, plus short turnaround time, no specific requirements for additional equipment or skilled technicians, all of these collectively contributed to its competence for mass testing. At the current stage, it cannot take the place of SARA-CoV-2 nucleic acid RT-PCR, but can be served as a complementary option for RT-PCR. The combination of RT-PCR and IgG-IgM combined test kit could provide further insight into SARS-CoV-2 infection diagnosis.

## Introduction

In December 2019, several cases of pneumonia of unknown etiology, now known as coronavirus disease 2019 (COVID-19), occurred in Wuhan, China, has been recognized as a pandemic by the World Health Organization (WHO) ^[1, 2]^. The disease has rapidly spread from Wuhan to other provinces in China and around the world. As of March 21, 2020, China has reported 81054 cases of confirmed COVID-19 and 3261 fatalities. Meanwhile, the number of confirmed COVID-19 and fatalities across the whole world are 267013 and 11201, respectively. The International Committee on Taxonomy of Viruses announced “severe acute respiratory syndrome coronavirus 2 (SARS-CoV-2)” as the name of the new virus caused COVID-19 ^[3]^.

Early detection, early reporting, early isolation, early diagnosis and early management have been proved to be useful in the course of COVID-19 patients ^[4]^. But how to detect early was an unmet demand in front of medical personnel. It is well known globally that virus nucleic acid high throughput sequencing or virus nucleic acid RT-PCR assay was the standard and recommended methods for detecting SARS-CoV-2, which caused COVID-19 pandemic across the whole world. High throughput sequencing can act as the ‘golden standard’ but it is highly restricted by its resource-dependent platform, especially in those resource-limited settings. Virus nucleic acid RT-PCR assay also suffer some limitations, long turnaround time, requirements for certified laboratories, skilled technicians and expensive equipment, and especially false negative results ^[5]^. Generally speaking, RT-PCT results can be affected by many factors, such as quality of samples, degeneration of virus RNA, and improper sampling of nasal and pharyngeal swab. A former study revealed that 5 cases who were continuously PCR negative but with chest CT scan positive results were finally PCR confirmed positive after a series of continuous RT-PCR testing, which will inevitably hinder COVID-19 pandemic control and limit the outbreak containment effort ^[6]^. This was in agreement with similar studies related to SARS in 2003, whose results showed that RT-PCR positive rate was only between 50% and 79% ^[7]^. In addition, the number and variety of diagnostic tests should be expanded based on the fact that severity and scope of the current COVID-19 situation around the globe necessitates greater testing capacity than is currently available. Therefore, detection methods that are faster and more convenient, and complementary to nucleic acid detection, are particularly important in the current epidemic prevention and control. Hence detecting antibodies of SARS-CoV-2 became a supplemented option for RT-PCR, no matter IgM or IgG or combined, which can provide another key piece of evidence for the diagnosis of viral infections.

The New Coronavirus Pneumonia Prevention and Control Program (7th edition) published by the National Health Commission of China also recommended that any suspicious cases together with positive IgM/IgG antibodies can be deemed as positive COVID-19 cases. However, data concentrating on its sensitivity, specificity were inadequate, which were of paramount value in the decision whether, when and how to use IgM/IgG antibodies detection. The aim of this study was to evaluate the sensitivity, specificity, accuracy, positive predictive value (PPV), negative predictive value (NPV), especially Kappa efficiency with PCR and its anti-interference ability.

## Method

### Patients

A total of 179 inpatient or outpatient COVID-19 cases, visited or admitted to General Hospital of Central Theatre Command between 1^st^ January 2020 and 12^th^ March 2020, were enrolled in this retrospective observational study. The selected 179 cases were divided into two groups based on the results of SARS-CoV-2 PCR results, namely PCR positive group and PCR negative group, which consisted of 90 cases and 89 cases, respectively.

PCR positive group consisted of 46 mild/common cases and 44 severe/critical cases while PCR negative group were composed of 5 clinical confirmed cases, 20 suspected cases and 64 cases, aged from 23 years to 80 years, who were diagnosed as other diseases other than COVID-19. The disease spectrums of 64 cases were as follows: 10 cases of Sjogren’s syndrome, 8 cases of diabetes, 6 cases of systemic lupus erythematosus, 5 cases of rheumatoid arthritis, 2 cases of dermatomyositis, 2 cases of connective tissue disease, 1 case of scleroderma, and 30 cases of common injuries with no underlying diseases. This study was approved by the Ethics Commission of General Hospital of Central Theatre Command.

### Sample collection

Approximate 5 ml fasting blood samples were drawn from all the enrolled participants and collected into serum separation hose and subsequently centrifuged at 2500 g for 10 min. The serum samples were used to detect. SARS-CoV-2 IgG/IgM antibodies. Corresponding nasal and pharyngeal swab specimens were also collected for SARS-CoV-2 RT-PCR.

### SARS-CoV-2 IgG/IgM antibody detection

The SARS-CoV-2 IgG/IgM antibody test kit is manufactured by a Chinese biotechnology company and has been cleared by China Food and Drug Administration. The protocols and routines of detection followed the guideline of this manufacture. It took about 15 minutes to finish the entire test. Three detection lines were on this kit. The Control (C) line appears when serum sample flowed through this stip. SARS-CoV-2 IgG/IgM will be indicated by a red test line in the G and M zone, respectively. There is one thing to mention, if C line does not appear, the test is invalid, and should be tested again with another completely new test kit.

### Data analysis

SPSS 20.0 was used for statistical analysis All tests were bilateral, and *p* less than 0.05 was considered statistically significant. Chi-square test or Fisher exact probability test was used to compare count data. Sensitivity, specificity, positive predictive value (PPV), negative predictive value (NPV) and Kappa efficiency were calculated.

## Results

### Example to show real testing results

Just as described in Method section, C line should appear in any test, otherwise the test is invalid. Either G or M or both lines appearing red lines shows presence of anti-SARS-CoV-2 IgG or anti-SARS-CoV-2 IgM or both antibodies in this detected sample. A representative picture for five patients blood testing results was shown in

**Figure 1.**
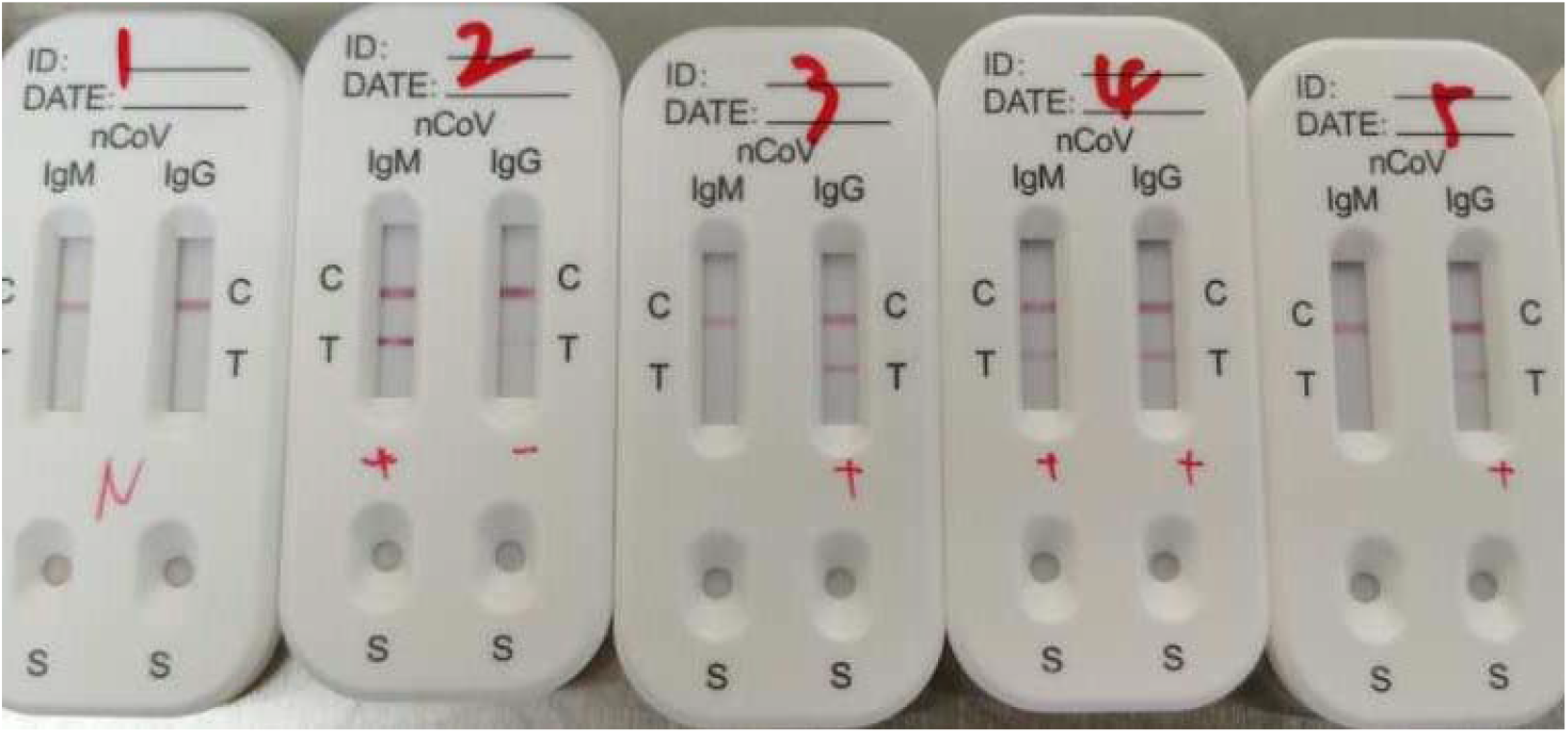
Result of Patient 1 was negative for both IgM and IgG antibodies. Result of Patient 2 was only IgM positive while results of patient 3 and 5 was only IgG positive. Result of patient 4 was positive for both IgM and IgG antibodies. Representative picture for five patients blood testing results. Result of Patient 1 was negative for both IgM and IgG antibodies. Result of Patient 2 was only IgM positive while results of patient 3 and 5 was only IgG positive. Result of patient 4 was positive for both IgM and IgG antibodies.

### Diagnostic indexes of SARS-CoV-2 IgG/IgM test kit

According to SARS-CoV-2 RT-PCR results, 179 patients under study were categorized as PCR positive group in 90 patients and PCR negative group in 89 patients. The average age (mean ± SD) in both groups were 76 ± 15 years and 56 ± 21 years, respectively. Time from illness onset to sample collection also significantly differed (30 ±17 VS 18 ±14, p < 0.001).

Of the 90 PCR positive samples, 77 were tested positive by SARS-CoV-2 IgG/IgM test kit, yielding a sensitivity of 85.6% (77/90). Meanwhile, of the 89 PCR negative sample, 8 samples were detected positive, resulting in a specificity of 91% (81/89). PPV, NPV and accuracy of this test kit was 95.1% (77/81), 82.7% (81/98), and 88.3% (158/179), respectively. Our study also calculated Kappa efficiency between SARS-CoV-2 IgG/IgM test kit and SARS-CoV-2 RT-PCR assay, with a result of 0.75 (p < 0.001), which further indicated that moderate agreement between these two methods.

### Accuracy of SARS-CoV-2 IgG/IgM test kit in different types of patients

PCR positive group was further divided into two subgroups: mild/common and severe/critical subgroup while PCR negative group was further divided into three subgroups: clinical confirmed, suspected cases and other disease subgroups (**Table 2**). Because PCR result in each subgroup was constant, diagnostic indexes except accuracy could not be calculated.

**Table 1.** Demographic information of enrolled participants and diagnostic indexes of SARS-CoV-2 IgG/IgM test kit

|  | PCR Positive | PCR Negative | p value |
| --- | --- | --- | --- |
| n | 90 | 89 | - |
| Age (Year) | 76 ± 15 | 56 ± 21 | < 0.001 |
| Sex (Female, %) | 30 (33.3%) | 51 (57.3%) | < 0.001 |
| Time from illness onset to sample collection (Days) | 30 ± 17 | 18 ± 14 | < 0.001 |
| Both IgG/IgM Positive | 32 | 5 | - |
| IgG Positive | 43 | 2 | - |
| IgM Positive | 2 | 1 | - |
| Diagnostic Indexes |  |  | - |
| Sensitivity | 85.6% (77/90) |  | - |
| Specificity |  | 91% (81/89) | - |
| PPV | 95.1% (77/81 ) |  | - |
| NPV | 82.7% (81/98) |  | - |
| Accuracy | 88.3% (158/179) |  | - |
| Kappa efficiency | 0.75 |  | < 0.001 |

Abbreviations: PPV: Positive predictive value; NPV: Negative predictive value.

**Table 2.**
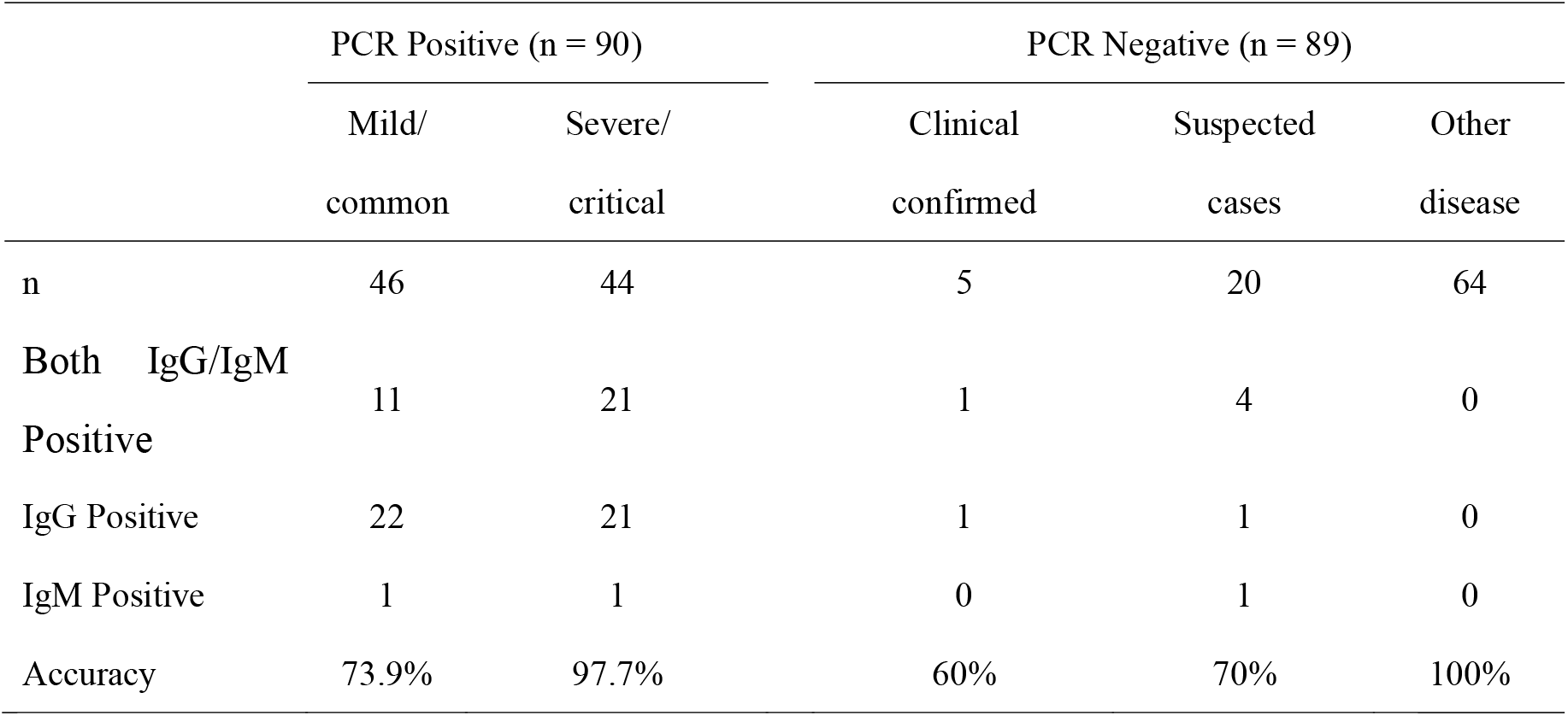
Accuracy in different subgroups.

|  | PCR Positive (n = 90) |  | PCR Negative (n = 89) |  |  |
| --- | --- | --- | --- | --- | --- |
|  | Mild/<br>common | Severe/<br>critical | Clinical<br>confirmed | Suspected<br>cases | Other<br>disease |
| n | 46 | 44 | 5 | 20 | 64 |
| Both IgG/IgM<br>Positive | 11 | 21 | 1 | 4 | 0 |
| IgG Positive | 22 | 21 | 1 | 1 | 0 |
| IgM Positive | 1 | 1 | 0 | 1 | 0 |
| Accuracy | 73.9% | 97.7% | 60% | 70% | 100% |

The highest accuracy was in other disease subgroup (100%). On the contrary, the lowest accuracy was in clinical confirmed subgroup (60%). The second highest accuracy was in severe/critical subgroup (97.7%) while the accuracy in the remained two subgroups was 73.9% in mild/common subgroup and 70% in suspected case subgroup, respectively.

### Diagnostic indexes of SARS-CoV-2 IgG/IgM test kit stratified by the time from illness onset to sample collection (days)

According to a previous study, IgM antibody could be detected in patient blood after 3 - 6 days of SARS infection while IgG antibody could be determined after 8 days of SARS infection ^[8]^. SARS-CoV-2 which caused COVID-19 belong to the same large family as that caused SARS ^[9]^, hence it is very important to calculate diagnostic indexes of this test kit stratified by the time from illness onset or infection time to sample collection.

A total of 115 inpatients, including 90 PCR confirmed positive cases, 5 clinical confirmed cases and 20 suspected cases, were divided into three groups according to the time from illness onset to sample collection: ‘0 – 7 days’ group, ‘8 – 15 days’ group and’ >= 16 days’ group (**Table 3**). The test kit under study performed the worst in ‘0 – 7 days’ group, generating a sensitivity of 18.8% and an accuracy of 40%. This test kit performed the best in ‘>= 16 days’ group, yielding a sensitivity of 100% together with an accuracy of 93.9%, though with a relatively low specificity (64.3%). In ‘8 – 15 days’ group, the diagnostic indexes of this test kit were 100% for sensitivity, 50% for specificity and 87.5% for accuracy, respectively.

**Table 3.** Diagnostic indexes of SARS-CoV-2 IgG/IgM test kit in different subgroups according to the time from illness onset to sample collection (days).

|  | 0 -7 days |  | 8 - 15 days |  | >= 16 days |  |
| --- | --- | --- | --- | --- | --- | --- |
|  | PCR + | PCR- | PCR+ | PCR- | PCR+ | PCR- |
| n | 16 | 9 | 6 | 2 | 68 | 14 |
| Both IgG/IgM Positive | 2 | 1 | 5 | 1 | 25 | 3 |
| IgG Positive | 0 | 1 | 0 | 0 | 43 | 1 |
| IgM Positive | 1 |  | 1 | 0 | 0 | 1 |
| Sensitivity | 18.8% |  | 100% |  | 100% |  |
| Specificity |  | 77.8% |  | 50% |  | 64.3% |
| Accuracy | 40% |  | 87.5% |  | 93.9% |  |

## Discussion

In the past week, COVID-19 has started behaving a lot like the once-in-a-century pathogen that international experts have been worried about^[10]^. Because the symptoms of COVID - 19 are not unique but similar to those of other diseases, testing is the only way to know for sure if someone is infected with the SARS-CoV-2 ^[11]^. Mass testing is therefore of paramount importance to curb SARS-CoV-2 epidemic. Virus nucleic acid RT-PCR is not suitable for large scale screening owing to the inherent properties. On the contrary, the qualitative detection of SARS-CoV-2 IgG/IgM antibodies in human serum, which is configured like a home pregnancy test, has the ability for mass testing.

As a rule of thumb, IgM, mainly found in the blood and lymph fluid, was the first antibody individual made against a new virus infection. It was detectable in patient’s blood after 3 – 6 days while IgG, the most abundant type of antibody, could be measured after 8 days. Positive IgM and IgG tests for SARA-CoV-2 antibodies detected in patient’s blood sample mean that it is likely that the individual became infected with SARA-CoV-2 recently or at the early stage of infection. If only the IgG is positive, then it is probable that the person had an infection sometime in the past or in the late stage of virus infection. Hence, in order to be applicable for different stages of COVID-19, combined detection of IgG and IgM antibodies were recommended.

The ease-of-use IgG/IgM combined test kit under study was developed based on gold immunochromatography assay (GICA), which can generate results in less than 15 minutes from human serum without laboratory equipment or skilled personnel or sample transportation. Its diagnostic indexes or so-called test characristics, including sensitivity, specificity, PPV, NPV and accuracy were evaluated to determine its diagnostic usefulness ^[11]^. Our results revealed that the above-mentioned five indexes were 85.6%, 91%,95.1%, 82.7%, and 88.3%, respectively which was in agreement with a recently published research ^[5]^. In that study, sensitivity and specificity were 88.66% and 90.63%. Sensitivity is the ability to diagnose those with the disease correctly identified as positive by the test. From the definition of sensitivity, we can suggest that manufacturer should try to improve IgG/IgM test kit’s detection sensitivity. Because the lower the sensitivity, the more false-negative cases. False-positive cases can be further confirmed by other detection methods; hence false-negative cases would pose more opportunities to infect people they contact.

Any person who got a positive IgG/IgM test result may want to know what the chance that he or she actually has the disease is. Our study further showed PPV of this IgG/IgM test kit was 95.1%, which indicated the proportion of people with a positive test result who actually have the disease was 95.1%. On the other hand, NPV of this test kit was just 82.7%, which demonstrated that the proportion of those with a negative result who do not have the disease was 82.7%. This result gave us a strong hint that negative IgG/IgM test results cannot exclude virus infection and repeat examination after one week or so was strongly suggested. Kappa efficiency was also investigated in our study (Kappa = 0.75, p < 0.001), which inflected moderate agreement between IgG/IgM test kit and RT-PCR assay.

Accuracy of SARS-CoV-2 IgG/IgM test kit in 5 clinical confirmed cases showed 2 out of the 5 cases were tested positive by IgG/IgM test kit while their RT-PCR results were negative (**Table 2**). SARS-CoV-2 infection starts at the lung not in the upper respiratory tract, hence sampling process has a large effect on final RT-PCR results ^[13]^, which might partially explain the high false-negative rate. However, this should not have any effect on SARS-CoV-2 IgG/IgM test kit because only venous blood was needed for this test. IgG/IgM test kit likely can remedy some false negatives inherent in respiratory swab samples and can be served as a complementary option to RT-PCR.

Compared with other similar studies, one of our strengths was that we recorded detailed time of each patient from illness onset to sample collection. From the results (**Table 3**), we can see that only 3 out of the 16 cases with positive PCR results in ‘0 – 7’ group were tested positive by SARA-CoV-2 IgG/IgM test kit, generating a sensitivity of just 18.8% (3/16). The time from illness onset to sample collection of these 16 patients in ‘0 – 7’ group was between 1 and 2 days. Patients in this group may be at the initial stage or ‘window period’ of SARS-CoV-2 infection, the concentration of antibodies was too low to be detected. In ‘8 – 15’ subgroup and ‘>= 16’ subgroup, the sensitivity of IgG/IgM test kit jumped from 18.8% to 100% whereas the specificity in these two groups were relatively low, 50% and 64.3%, respectively. It was very difficult to explain, because we were unsure the low specificity was caused by false negative results of RT-PCR, taking high false negative rate of RT-PCR into consideration, or by false positive results of IgG/IgM test kit itself or by small sample size. The other strength of our study was enrollment of 34 patients diagnosed with autoimmune disease, such as Sjogren’s syndrome, systemic lupus erythematosus, rheumatoid arthritis, and connective tissue disease. The enrollment of these patients was to determine IgG/IgM test kit’s anti-interference ability. Our results showed IgG/IgM test kit performed well in these patients.

However, some limitations also should be pointed out. First, due to the limited time, diagnostic indexes were only evaluated in serum, but not other blood sample types, such as fingertip blood and plasma. Second, small sample size should also be taken into consideration.

## Conclusion

The sensitivity and specificity of this ease-of-use IgG/IgM combined test kit were good, plus short turnaround time, no specific requirements for additional equipment or skilled technicians, all of these can collectively contributed to its competence for mass testing. At the current stage, it cannot take the place of SARA-CoV-2 nucleic acid RT-PCR, but can be served as a complementary option for RT-PCR. The combination of RT-PCR and IgG/IgM combined test kit could provide further insight into SARS-CoV-2 infection diagnosis.

## Data Availability

none

## References

[1] Phelan A L, Katz R, Gostin L O. The Novel Coronavirus Originating in Wuhan, China: Challenges for Global Health Governance[J]. JAMA,2020.

[2] Bedford J, Enria D, Giesecke J, et al. COVID-19: towards controlling of a pandemic[J]. Lancet,2020.

[3] Andersen K G, Rambaut A, Lipkin W I, et al. The proximal origin of SARS-CoV-2[J]. Nature Medicine,2020.

[4] Guan W, Ni Z, Hu Y, et al. Clinical Characteristics of Coronavirus Disease 2019 in China[J]. The New England journal of medicine,2020.

[5] Li Z, Yi Y, Luo X, et al. Development and Clinical Application of A Rapid IgM-IgG Combined Antibody Test for SARS-CoV-2 Infection Diagnosis[J]. Journal of Medical Virology,2020.

[6] Xie X, Zhong Z, Zhao W, et al. Chest CT for Typical 2019-nCoV Pneumonia: Relationship to Negative RT-PCR Testing[J]. Radiology,2020:200343.

[7] Yam W C, Chan K H, Poon L L, et al. Evaluation of reverse transcription-PCR assays for rapid diagnosis of severe acute respiratory syndrome associated with a novel coronavirus[J]. J Clin Microbiol,2003,41(10):4521–4524.

[8] Lee H K, Lee B H, Seok S H, et al. Production of specific antibodies against SARS-coronavirus nucleocapsid protein without cross reactivity with human coronaviruses 229E and OC43[J]. J Vet Sci,2010,11(2):165–167.

[9] Gralinski L E, Menachery V D. Return of the Coronavirus: 2019-nCoV[J]. Viruses,2020,12(2):135.

[10] Gates B. Responding to Covid-19 - A Once-in-a-Century Pandemic?[J]. The New England journal of medicine,2020.

[11] Wang D, Hu B, Hu C, et al. Clinical Characteristics of 138 Hospitalized Patients With 2019 Novel Coronavirus–Infected Pneumonia in Wuhan, China[J]. JAMA,2020.

[12] Rao G. Remembering the meanings of sensitivity, specificity, and predictive values[J]. The Journal of family practice,2004,53(1):53.

[13] Huang C, Wang Y, Li X, et al. Clinical features of patients infected with 2019 novel coronavirus in Wuhan, China[J]. The Lancet,2020.

